# Mast cells are upregulated in Hidradenitis Suppurativa Tissue, associated with epithelialized tunnels and normalised by Spleen Tyrosine Kinase Antagonism

**DOI:** 10.1101/2023.04.02.23287928

**Authors:** A Flora, R Jepsen, EK Kozera, JA Woods, GD Cains, M Radzieta, SO Jensen, M Malone, JW Frew

## Abstract

Mast cells have traditionally been associated with allergic inflammatory responses; however they play important roles in cutaneous innate immunity and wound healing. The Hidradenitis Suppurativa tissue transcriptome is associated with alterations in innate immunity and wound healing associated pathways, however the role of Mast cells in the disease is unexplored. We demonstrate that Mast cell associated gene expression (using whole tissue RNAseq) is upregulated, and in-silico cellular deconvolution identifies activated mast cells upregulated and resting mast cells downregulated in lesional tissue. Tryptase/Chymase positive Mast cells (identified using IHC) localize adjacent to epithelialised tunnels, fibrotic regions of the dermis and at perivascular sites associated with Neutrophil Extracellular Trap formation and TNF-alpha production.

Treatment with Spleen Tyrosine Kinase antagonist (Fostamatinib) reduces the expression of mast cell associated gene transcripts, associated biochemical pathways, and number of tryptase/chymase positive mast cells in lesional hidradenitis suppurativa tissue.

This data indicates that although Mast cells are not the most abundant cell type in Hidradenitis Suppurativa tissue, the dysregulation of mast cells is associated with B cell/plasma cell inflammation, inflammatory epithelialized tunnels and epithelial budding. This provides an explanation as to the mixed inflammatory activation signature seen in HS, the correlation with dysregulated wound healing and potential pathways involved in the development of epithelialized tunnels.

## INTRODUCTION

Hidradenitis Suppurativa (HS) is a chronic inflammatory skin disease characterised by panful inflammatory nodules, deep-seated abscesses and chronically draining epithelialized tunnels with a predilection with flexural skin^1,2^. The inflammatory transcriptome of tissue in HS includes a combination of Th17 dysregulation, B cell, plasma cell and immunoglobulin production as well as transepithelial neutrophil migration in epithelialized tunnels^3-6^. The precise pathogenesis of the disease is incompletely understood, although features of HS have known overlaps with other diseases including Psoriasis Vulgaris^7^, impaired wound healing^3^, as well as the fibroinflammatory features of diseases such as Scleroderma and Rheumatoid Arthritis^8,9^.

Both impaired wound healing and fibroinflammatory disorders illustrate dysregulated mast cell function, cross talk with B cells and interaction with fibroblasts^10,11^. These complex interactions lead to further activation of pro-inflammatory pathways and production of cytokines including IL-1B, IL-6 and IL-8^10-15^. Mast cells are associated with cathelicidin production in rosacea^15^ and play important roles in impaired cutaneous epithelialization and hypertrophic scar formation.^11-14,16^ Both of these characteristics are central features in HS. Mast cells have been identified as upregulated in HS lesional skin as well as associated with pruritus in the disease^17,18^.

Whilst immunoglobulin E (IgE) and histamine are the most well-recognised triggers and products of mast cells respectively, mast cells are capable of responding to microbiological triggers through lectin binding receptors and direct production of a wide array of pro-inflammatory cytokines including TNF-a^14,15,19-21^. Alterations in lectin binding are associated with recurrent infections, autoinflammatory disease and have been reported coexistent with HS which suggest a potential role for non-IgE mast cell pathways as contributory in HS. Novel therapeutic targets in HS have focused upon targeting B cells as well as small molecules associated with multiple inflammatory pathways. One of these potential small molecules of interest is spleen tyrosine kinase (SYK), associated with mast cells, B cells, plasma cells, and cells of the monocyte lineage^19-21^.

In this study we aimed to assess the presence and characteristics of Mast cells in HS lesional tissue compared to non-lesional tissue and healthy controls. Additionally, the impact of SYK antagonism with Fostamatinib (as part of the Phase 2 clinical trial of Fostamatinib in HS NCT05040698) upon the presence and activity of mast cells was assessed using gene expression profiles and immunohistochemical markers in HS lesional tissue.

## MATERIALS AND METHODS

### Patients and Tissue Collection

This study was approved by the human research ethics committee of Sydney South West Area Health Service (Sydney Australia). A total of 20 participants underwent lesional and non lesional biopsies (6mm in diameter) based upon previously published definitions^24,25^. All. Biopsies were immediately stored in RNA Later and frozen at -80C until processing. Standard wound care was undertaken for each of the biopsy sites. All patients were not on any active therapy at the time of biopsies. The clinical characteristics of participants are included in Supplementary Data.

### Skin Biopsy mRNA Quantification

Total RNA was extracted using the Qiagen RNeasy kit as per standard protocol (Qiagen, Valencia, CA, USA). RNA was sequenced using the Novaseq S4 (Illumina, San Diego, CA, USA) Differential expression was estimated with DESeq2 and VST to log_2_ transform the normalized counts. Differentially expressed genes were defined as fold change (FCH) of ≥ ¦1·5¦ and false discovery rate⍰≤⍰0·05.

### Pathway Analysis

Enrichment analyses were completed using Protein ANalysis THrough Evolutionary Relationships (PANTHER) classification system (www.pantherdb.org). Analyses were performed using whole tissue RNAseq data (Figure 1) as well as for Nanostring Gene Expression Panels. Nanostring gene expression panels included Baseline (prior to fostamatinib administration) and Week 4 of fostamatinib therapy lesional tissue samples (Figure 5).

**Figure 1:**
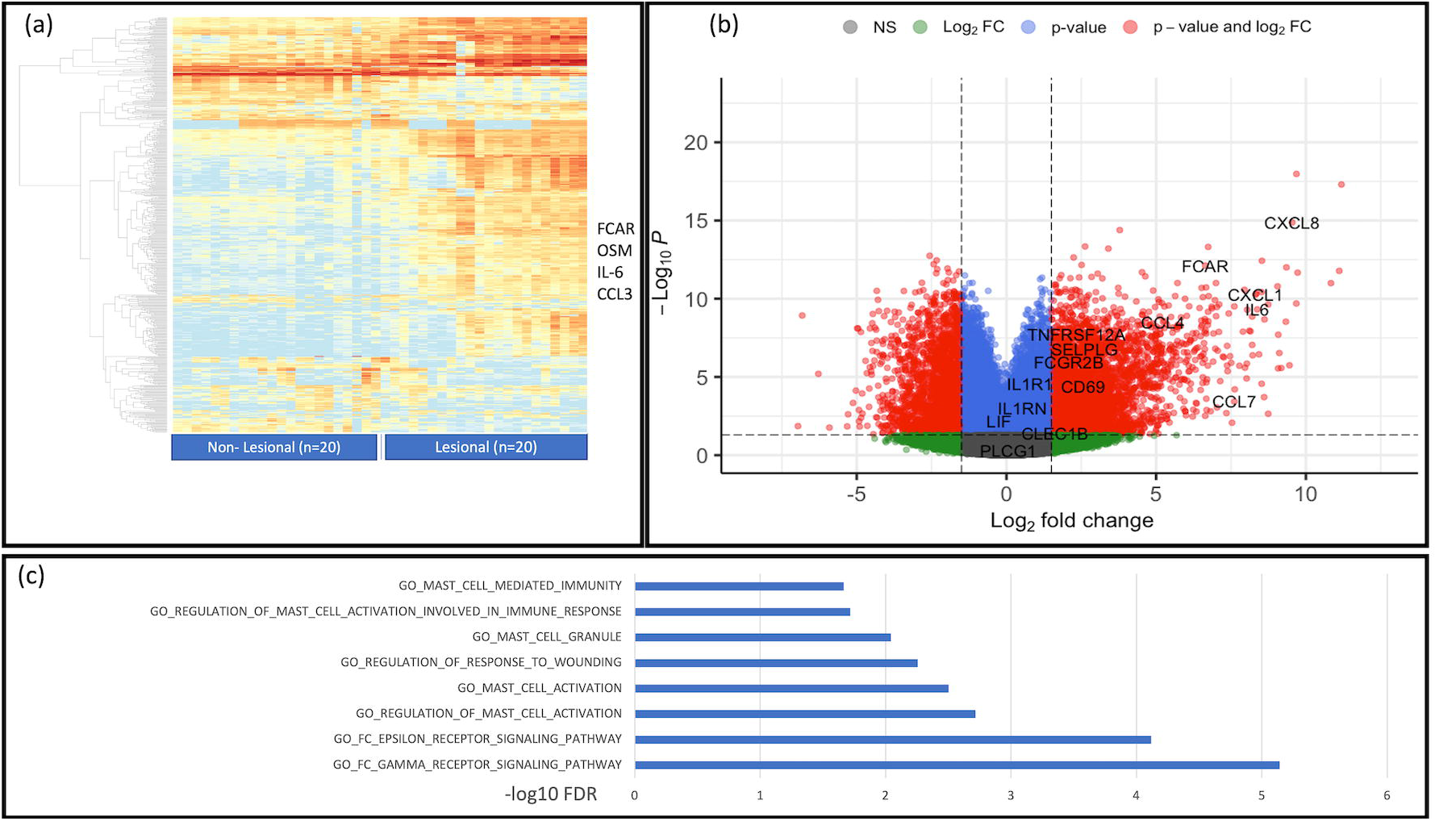
Whole Tissue RNAseq demonstrated upregulation of genes associated with Mast cell activity in lesional tissue of HS compared to non-lesional tissue. (a): Heatmap demonstrating overall tissue transcriptome with highest DEG pertaining to B cell and keratinocyte related genes-Cytokines and proteins associated with Mast cell activity also dyseregulated. (B): Volcano plot demonstrating the statistical significance of some mast cell associated genes which also overlap with possible other innate inflammatory activity

### CIBERSORTx

In silico deconvolution was undertaken using the CIBERSORTx web application ((https://cibersortx.stanford.edu/index.php) in order to estimate the abundance of immune cell and mast cell subpopulations in baseline lesional and non-lesional tissue using whole tissue RNAseq. The utilized immune cell reference matrix was based upon previously published human skin scRNAseq data (GSE65136) with stratification into immune cell subsets a previously defined.

### Nanostring Analysis

Extracted RNA was analysed using the NCounter system (Nanostring) using the Human Fibrosis V2.0 gene panel. Raw data was processed using the Nanostring Nsolver (version 4.0.70) analysis software using quality control and normalization procedures derived from the NormqPCR R package as previously described^26^. Differentially expressed genes (DEGs) were defined as >1.5 Log2Fold change with a false discovery rate<0.05 and p value<0.05.

### Immunohistochemistry

IHC staining was undertaken using primary antibodies listed in Supplementary Files. Stained slides underwent semiquantitative analysis using standardized, previously published methods^27^. Differences between groups were analyzed using the Wilcoxon rank sum test for two variables and/or omnibus test using one-way ANOVA with adjustment for multiple comparisons was made using the Benjamini Hochberg procedure.

### Statistical Analysis

RNA sequencing statistical analysis was performed in R language (www.r-project.org; R Foundation, Vienna, Austria) using publicly available packages (www.bioconductor.org). Means of each group and differences between groups were estimated using least-square means. Hypothesis testing was undertaken using the general framework for linear mixed-effect models of the limma package. tissue was considered a fixed factors while random effect related to subjects was included. Unsupervised clustering was performed with the Euclidean distance and average agglomeration. RNAseq data P-values were corrected for multiplicity using Benjamini– Hochberg procedure.

## RESULTS

### RNA sequencing demonstrates upregulation of Mast cell associated gene signatures including FCAR, CCL3, OSM as well as Mast cell associated pathways

Whole tissue RNAseq from lesional and non-lesional tissue from 20 individuals with HS (clinical and demographic data of included participants are included in Supplemental Data) identified differentially expressed genes associated with mast cell activity including *FCAR, CCL4, CCL7, CCL3, CXCL1* and *OSM* (Figure 1a,b). Pathway analysis demonstrated positive enrichment of pathways associated with regulation of mast cell activation, FC epsilon receptor signaling and regulation of response to wounding (Figure 1c). A full list of differentially expressed genes is included in Supplementary Data.

### In Silico Deconvolution Analysis Identifies Significant Increases in activated mast cells in lesional tissue of HS compared to non-lesional skin

Using CibersortX, in silico deconvolution of whole tissue RNAseq was used to identify transcriptomic signatures indicative of mast cell activation (Figure 2). Whilst no significant difference in overall mast cell proportions was identified, the proportion of resting mast cells was significantly decreased in lesional tissue compared with non-lesional tissue (Figure 2b). Additionally, the proportion of activated mast cells were significantly elevated in lesional tissue compared to non lesional tissue (Figure 2b).

**Figure 2:**
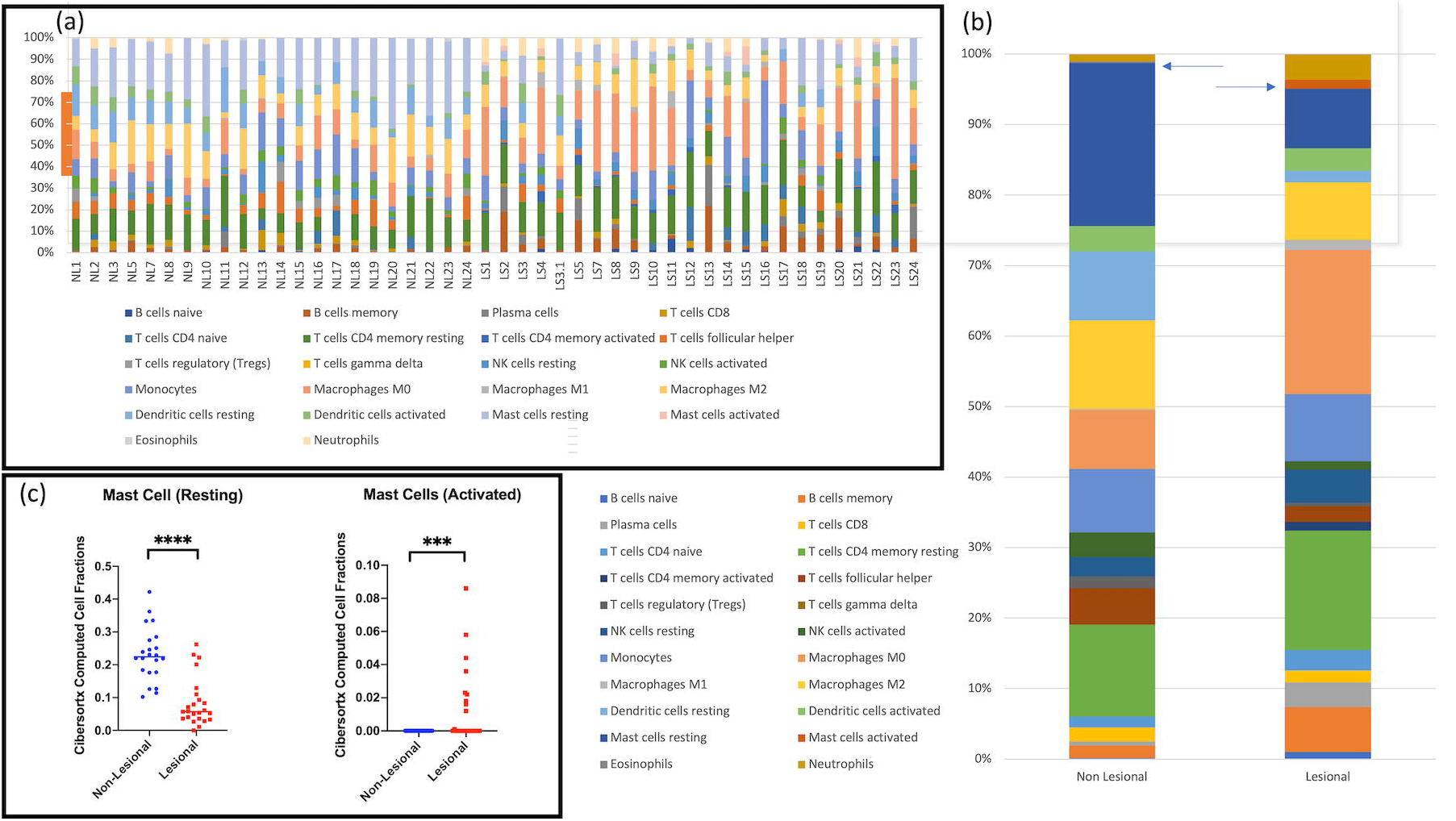
CibersortX In Silico Deconvolution Analysis Identifies Significant Increases in Mast cell and activated mast cells in lesional tissue of HS. (A): Individual deconvolution significant expansion of B cell, macrophage and CD4 T cell populations in lesional versus non lesional tissue. (B) but also a statistically significant decrease in resting mast cell and increase in activated mast cell populations (C).

### Immunohistochemistry identifies Mast cell clustering around folliculosebaceous units, and epidermal budding with significantly increased numbers of mast cells in lesional and peri-lesional tissue

Immunohistochemical (IHC) staining for mast cells validates the results seen in RNA sequencing and CIbersortX in-silico deconvolution analysis. IHC staining identified presence of mast cells adjacent to pilosebaceous units (Figure 3a) in lesional, perilesional and non-lesional tissue. Increased numbers of mast cells were identified alongside epithelial buds and active draining epithelialised tunnels (Figure 3b). Quantification of mast cell numbers demonstrated a statistically significant difference between lesional, perilesional and non-lesional tissue (Figure 3c). IHC staining with mast cell tryptase (Figure 3d) and mast cell chymase (Figure 3e) also demonstrated significant differences between the number of positive staining cells between lesional, perilesional and non-lesional tissue. Staining of serial sections identified mast cells expressing both mast cell tryptase and mast cell chymase (M_CT_). Expression was present both in epidermal, dermal sections as well as surrounding epithelialised tunnels and tunnel-associated vasculature (Figure 4 and Supplementary Figure 1). M_CT_ cells were significantly more abundant in Hurley stage 3 patients compared to hurley stage 2 patients (Figure 4) as well as those patients with epithelialised tunnels compared to those without (Figure 4). Mast cells colocalized in the deep dermis surrounding vasculature with CD20+ B cells and CD138+ plasma cells as well as MPO+ neutrophils (Supplementary FIgure 1).

**Figure 3:**
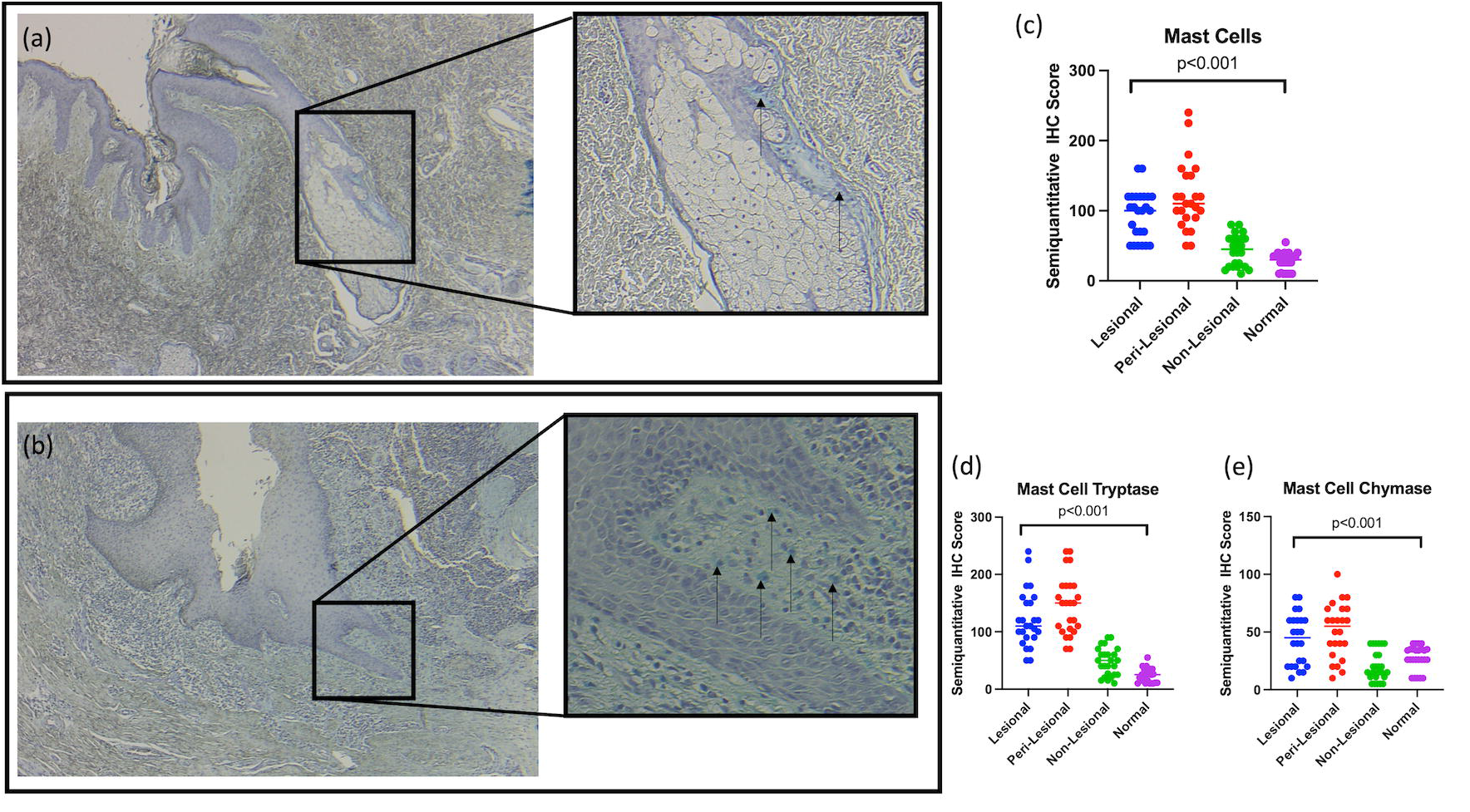
Immunohistochemistry staining for Mast cells in Leisonal and Non Lesional tissue demonstrating normal clustering around hair follicles, sebaceous glands in non-lesional unaffected tissue, with increased numbers of Mast cells around epithelial buds and active draining epithelialized tunnels.

**Figure 4:**
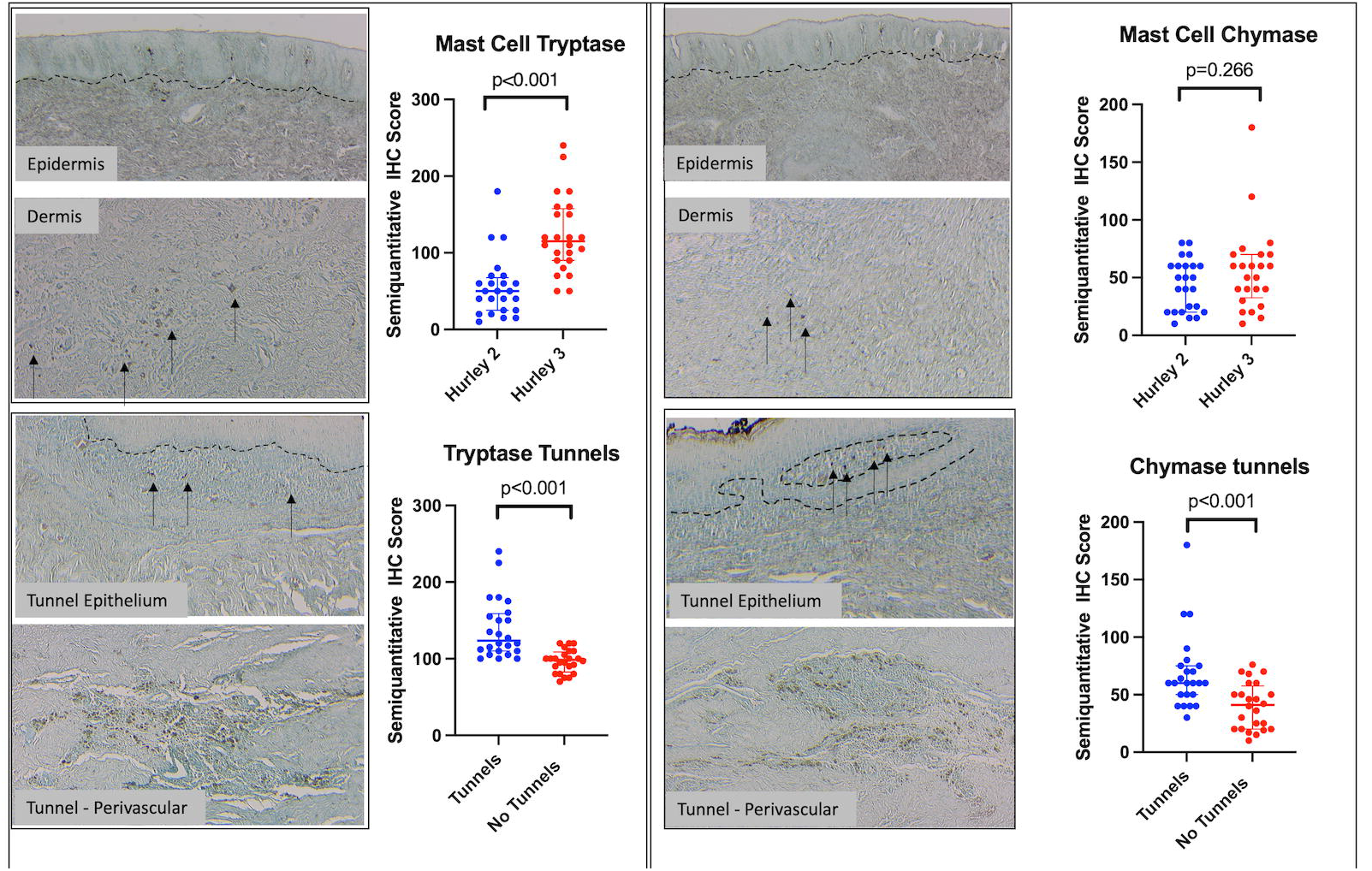
Immunohistochemical staining for mast cell tryptase (a) and mast cell chymase (b) demonstrating increased numbers surrounding vasculature and tunnel epithelium, with numerically increased numbers in Hurley stage 3 patients compared to Hurley stage 2 patients, and biopsy specimens with epithelialised tunnels as opposed to without epithelialised tunnels.

**Figure 5:**
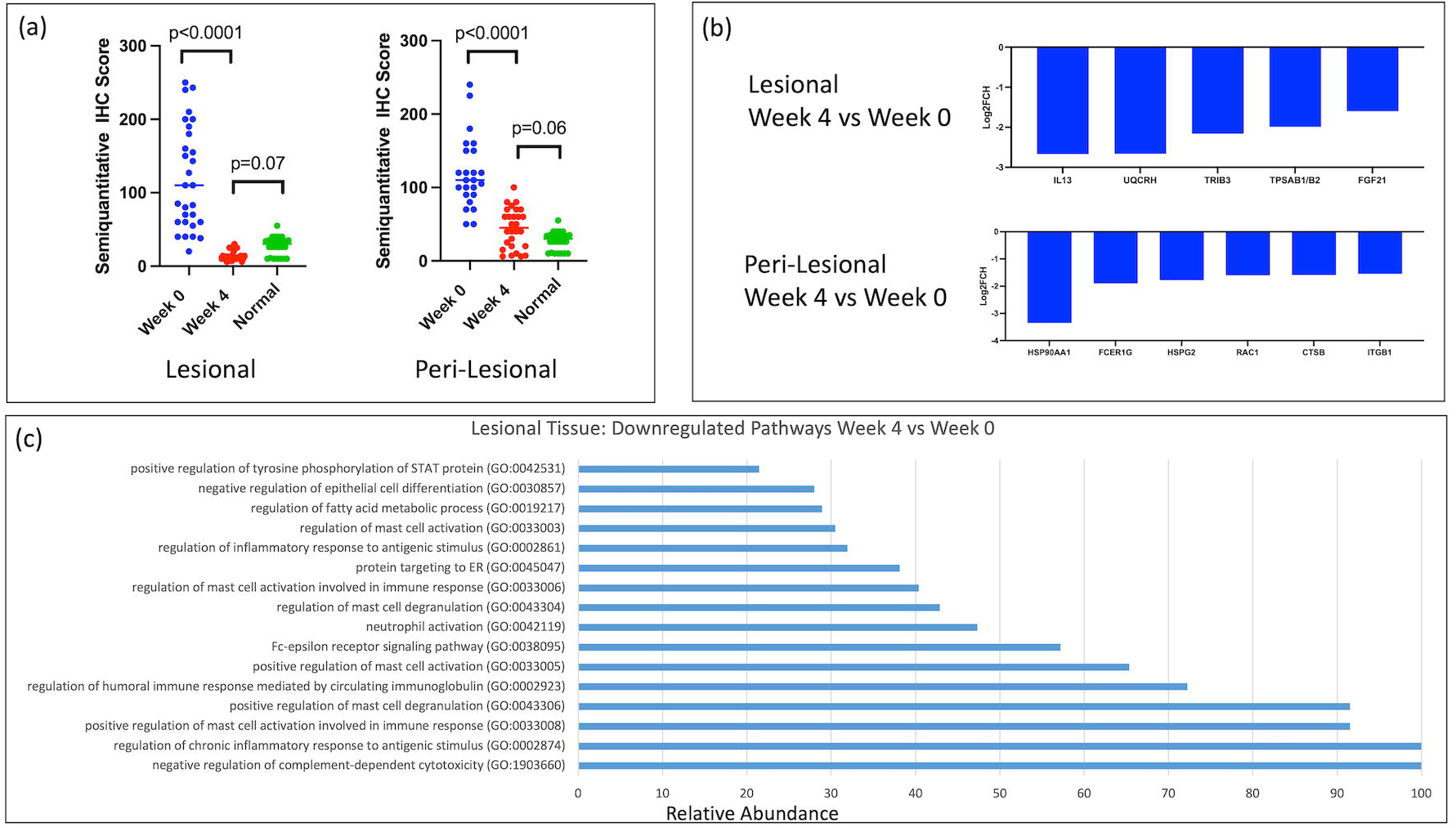
(a) Semiquantitative immunohistochemical evaluation of skin biopsies at Week 0 and Week 4 after administration of 100mg BID of Fostamatinib demonstrating a significant reduction in mast cell numbers. (b) Gene expression profiling with Nanostring demonstrating significant reduction in Mast cell associated gene expression at Week 4 compared to Week 0. (c) Pathway analysis demonstrating reduction in mast cell associated pathways at Week 4 compared to baseline.

### Treatment with Spleen Tyrosine Kinase Antagonist Fostamatinib normalized Mast cell populations in lesional HS tissue, with decrease in Mast cell associated gene expression and pathways compared to baseline

Spleen Tyrosine Kinase (SYK) inhibition with Fostamatinib at a dose of 100mg twice daily significantly reduced the presence of mast cells as measured by semiquantitative IHC (Figure 5a). The level of mast cells at week 4 of fostamatinib therapy was comparable to that in healthy controls and this effect was seen in both lesional tissue and perilesional tissue. Significant modulation of gene expression as measured by the Human Fibrosis V2.0 Nanostring gene expression assay was also noted at Week 4 compared to baseline (Figure 5b). Genes including *IL13, TRIB3, TSPAB1/2, FCER1G* and *RAC1* (involved with mast cell sensitisation and signalling) were significant downregulated at week 4 compared with baseline. Pathway analysis demonstrated high levels of enrichment associated with mast cell activation, mast cell degranulation and Fc-epsilon receptor signalling. (Figure 5c).

## DISCUSSION

This study presents novel gene expression and immunohistochemical data pertaining to the presence of activated mast cells in lesional and perilesional tissue of Hidradenitis Suppurativa. In comparison to healthy controls and non-lesional tissue, mast cells are increased in number and activated mast cells are significantly upregulated in lesional compared to non-lesional tissue. Within biopsy specimens, mast cells are identified in areas surrounding the pilosebaceous unit, at the advancing edge of epithelial buds in the epidermis as well as to areas surrounding epithelialised dermal tunnels. They also colocalise with B cells, plasma cells and neutrophils in the deep dermis surrounding vascular structures. This raises the prospect that mast cell activation may be involved with epithelial mesenchymal transition (EMT) which has been identified as present in HS lesions with epithelialised tunnels^28,29^. The activity of mast cells in HS tissue has previously been described^17,18^, however this study provides a much higher degree of resolution as to the localisation of mast cells as well as associations with clinical characteristics such as Hurley Staging and presence of epithelialised tunnels. The presence of mast cells may help explain pruritus in the disease, which is a largely underappreciated symptom of HS^18^.

RNA sequencing has illustrated similarities between the HS transcriptome and wound healing^3^, suggesting aberration in wound healing responses are involved in the pathogenesis of disease. Mast cells contribute to wound healing through EMT mechanisms, vascular dilatation and fibrinogenesis through release of various cytokines and chemokines^31^. Upregulation of S100 proteins and downregulation of Dermicidin are prominent features in the transcriptome of HS^3^. Evidence from in vitro studies have associated alterations in these products with variations in mast cell function and IL-6, IL-8 and TGF-B production^31^. This evidence, along with the localisation of mast cells at discrete areas in HS tissue suggest that their role may be less associated traditional allergic histamine responses and more associated with EMT, pro-fibrotic processes and influx of other inflammatory cells such as B cells, plasma cells and neutrophils^31^.

Mast cells are associated with fibrosis in various organ systems including the skin through production of TGF-B, IL-1, IL-33 and IL-8^14,15^. They also demonstrate interactions with other inflammatory cells including B cells and modulate neutrophil influx from vasculature into tissue^15^. The colocalization of mast cells in HS patient with Hurley 3 disease and epithelialised tunnels, as well as HS tissue in areas associated with EMT, B cells and neutrophil activity all support the role of mast cells as supportive players in the complex inflammatory milieu of HS^32^. SYK antagonism with Fostamatinib demonstrated significant modulation of mast cell numbers and mast cell associated gene expression after four weeks of therapy. This, alongside the clinical efficacy demonstrated in our Phase 2 proof-of-concept study (NCT05040698) support the role of small molecular targets modulating multiple aspects of innate and humoral immunity in the treatment of HS. Future analysis of this clinical trial data will enable deeper investigation into the precise mechanisms of SYK inhibition in HS at higher doses and further timepoints.

## Data Availability

Baseline RNA sequencing Data is publicly available using SRA Bioproject ID: PRJNA912704
Nanostring Data is publicly available using GEO Accession Number GSE220454

## Data Availability Statement

Baseline RNA sequencing Data is publicly available using SRA Bioproject ID: PRJNA912704 Nanostring Data is publicly available using GEO Accession Number GSE220454

## Acknowledgements

Nil

## Author Contributions

Akshay Flora: Investigation, Methodology, Visualization, Writing-Revision

Rebecca Jepsen: Investigation, Methodology, Writing-Revision

Emily Kozera: Methodology, Writing-Revision

Jane Woods: Methodoogy, Writing-Revision

Geoff Cains: Methodology, Writing-Revision

Michael Radzieta: Investigation Data Curation, Formal Analysis, Methodology, Writing-Revision

Matthew Malone: Investigation, Data Curation, Methodology, Writing-Revision

John Frew: Conceptualization, Data Curation, Formal Analysis, Investigation, Methodology, Software, Visualization, Writing-Original Draft Preparation

## TABLES AND FIGURE LEGENDS

Supplementary Figure 1: Individual patient cell proportion data from CibersortX in-silico deconvolution examining mast cells in lesional and non-lesional tissue of Hidradenitis Suppurativa.

Supplementary Figure 2: Additional immunohistochemical staining demonstrating clustering of tryptase and chymase positive mast cells with CD20+ B cells, CD138+ plasma cells and MPO+ neutrophils surrounding dermal vasculature.

Supplementary Table 1: Patient Demographics and Disease Characteristics

Supplementary Table 2: Differentially Expression Genes in whole tissue RNAseq

Supplementary Table 3: Immunohistochemical Antibodies used in this study.

Supplementary Table 4: Differentially Expressed Genes in Nanostring Gene Expression Panel.

